# Predicting missed health care visits during the COVID-19 pandemic using machine learning methods: Evidence from 55,500 individuals from 28 European Countries

**DOI:** 10.1101/2022.03.01.22271611

**Authors:** Anna Reuter, Šime Smolić, Till Bärnighausen, Nikkil Sudharsanan

## Abstract

**Background:** The COVID-19 pandemic has led many individuals to miss essential care. Machine-learning models that predict which patients are at greatest risk of missing care visits can help health administrators prioritize retentions efforts towards patients with the most need. Such approaches may be especially useful for efficiently targeting interventions for health systems overburdened by the COVID-19 pandemic.

**Methods:** We compare the performance of four machine learning algorithms to predict missed health care visits based on common patient characteristics available to most health care providers. We use data from 55,500 respondents of the Survey of Health, Ageing and Retirement in Europe (SHARE) COVID-19 survey (June – September 2020) in conjunction with longitudinal data from waves 1-8 (April 2004 – March 2020). We use stepwise selection, group lasso, random forest and neural network algorithms and employ 5-fold cross-validation to test the prediction accuracy, sensitivity, and specificity of the selected models.

**Findings:** Within our sample, 15.5% of the respondents reported any missed essential health care visit due to the COVID-19 pandemic. All four machine learning methods perform similarly in their predictive power. When classifying all individuals with a predicted probability for missed care above 17% as at risk of a missed visit, they correctly identify between 41% and 53% of the respondents at risk, while correctly identifying between 74% and 64% of the individuals not at risk. We find that the sensitivity and specificity of the models are strongly related to the risk threshold used to classify individuals; thus, the models can be calibrated depending on users’ resource constraints and targeting approach. All models had an area under the curve around 0.62, indicating that they outperform random prediction.

**Interpretation:** Pandemics such as COVID-19 require rapid and efficient responses to reduce disruptions in health care. Based on characteristics available to health insurance providers, machine learning algorithms can be used to efficiently target efforts to reduce missed essential care.

**Funding:** Research in this article is a part of the European Union’s H2020 SHARE-COVID19 project (Grant Agreement No. 101015924).

## Background

Pandemics disrupt regular health care delivery. During the first COVID-19 wave in Europe, primary care visits, hospital admissions, and emergency department visits all declined substantially, already before lockdowns were imposed.^1–4^ Even hospital admissions for serious acute health conditions such as heart failure and myocardial infarction decreased considerably.^5,6^ These care disruptions are thought to be a main reason why cardiovascular disease mortality increased in the early stages of the pandemic.^7,8^ Disruptions to essential care may also have longer term effects that persist beyond the pandemic.^9^ In Germany, for example, diagnoses of diabetes, dementia, depression and stroke decreased to a larger extent than the number of physician consultations,^4^ indicating that those in need of preventive care delayed essential health care visits.

The COVID-19 pandemic is the most recent example of pandemics disrupting health care. Similar declines in health care usage were observed during the outbreak of SARS in Taiwan in 2003 or MERS in South Korea in 2015.^10–13^ In contrast to these previous pandemics, however, COVID-19 spans across the globe and is characterized by a longer duration with multiple waves. Thus, while the utilization of health care services recovered quickly in the case of SARS and MERS,^13,14^ current evidence indicates a slow recovery and repeated disruption of health care visits during COVID-19.^1,3,6^ The relentless nature of the COVID-19 thus carries greater risks than prior pandemics. Therefore, individuals may end up chronically forgoing essential care or delaying necessary care so long that they increase their risk of severe health complications.

Individuals at risk for care disruptions need to be efficiently contacted and reconnected to health services to prevent them from forgoing or delaying essential care due to the COVID-19 pandemic. Hospital administrators and health insurance providers are particularly well placed for this role as they can contact patients quickly and at a large scale. However, they face two important challenges. First, targeting efforts need to be accurate and straightforward without requiring substantial additional resources to not overwhelm the limited capacities of health systems already under pressure. The health-system strain created by the COVID-19 pandemic likely results in limited capacity to contact all patients; instead, health administrators and insurance providers need a way of filtering patients by their risk of care disruptions and contacting those with the greatest needs. A second challenge is that while health administrators and insurance providers hold information on health care visits, they usually cannot observe whether they were postponed or cancelled and why. Building prediction models using existing large-scale survey data has the potential to address both of these gaps. Survey data contain information on patients and whether they missed visits. This allows for building models that predict the risk of missed visits using the patient characteristics available to health administrators and insurance providers. These models can then be applied by health administrators and insurance providers to existing patient information to predict each patient’s risk of missed visits, and based on this predicted risk, target efforts to those who would benefit the most.

In this study, we use data from a special wave of the Survey of Health, Ageing and Retirement in Europe (SHARE) to build and evaluate models that use common patient characteristics to predict missed health care visits. We compare the performance of four popular machine learning algorithms to determine if some algorithms are better suited to certain targeting approaches and resource constraints compared to others. Our findings have immediate and long-term relevance. The models we present here can be used by health administrators and insurance providers to target individuals with efforts to encourage continuity of care for current and future waves of the COVID-19 and other future pandemics.

## Methods

### Sample

We use data from the Survey of Health, Ageing and Retirement in Europe (SHARE), waves 1-8 and the first wave of the COVID-19 survey.^15–24^ The SHARE panel includes health and socioeconomic data of respondents aged 50 or older and their partners, and covers the European Union (except Ireland), Switzerland and Israel.^15^ While the in-person data collection for wave 8 had to be stopped due to the outbreak of COVID-19 in March 2020, computer-assisted telephone interviews were conducted between June and August 2020 for a special survey on COVID-19, covering 57,303 individuals. The SHARE study was approved by the Ethics Committee at the University of Mannheim (waves 1‐4) and by the Ethics Council of the Max‐Planck‐Society (waves 5‐8). Additionally, country-specific ethics committees or institutional review boards approved implementations of SHARE in the participating countries. All study participants provided informed consent.

### Missed health care visits

Our primary outcome is whether an individual did not attend an essential health care visit. The SHARE COVID-19 questionnaire asked individuals whether they experienced three types of missed health care visits due to COVID-19: (1) forgone medical treatment due to fear of becoming infected; (2) postponement of scheduled treatments by the health provider due to COVID-19; (3) denied appointment since the outbreak of the pandemic. Our main analyses consider any reported missed health care visit; secondarily, we also examine the different reasons for missed health care visits separately. We excluded missed visits to dentists and specialists (one mutual answer category in the questionnaire) as we deem it likely that missed visits in this category are mainly driven by postponed dentist check-ups.

### Predictors

We focus our analysis on possible predictors of missed health care visits that health insurance providers and hospital administrators are likely to have access to. This includes age, sex, education, latest reported employment status, past diagnoses, and past medication use. We defined past diagnoses and medication as the respondent stating in any of the past SHARE waves that a certain condition was (ever) diagnosed/a certain medication was taken. We dropped diagnoses and medications that were asked for irregularly across SHARE waves (asthma, arthritis, osteoporosis, benign tumor, chronic kidney disease, drugs for asthma, drugs for osteoporosis, drugs for suppressing inflammation) to circumvent sample selection problems. Except for age, all predictors are categorical. Sex is measured with an indicator for being female (the data distinguishes between male and female), education with six distinct categories (none, primary, lower secondary, upper secondary, post-secondary, and tertiary education), and employment with six categories as well (retired, working, unemployed, disabled, homemaker, other).

### Missingness

This was a complete case analysis. Of the 57,303 individuals interviewed in the SHARE COVID-19 wave, we excluded 1,799 individuals (3% of the eligible sample) that had missing data on either the outcome or predictors (Figure 1). This resulted in a sample of 55,504 individuals (97% of the eligible sample). We additionally excluded four randomly selected individuals to enable an even split into five samples for cross-validation procedures (more details below). All data preparation was conducted in Stata 17.

**Figure 1.**
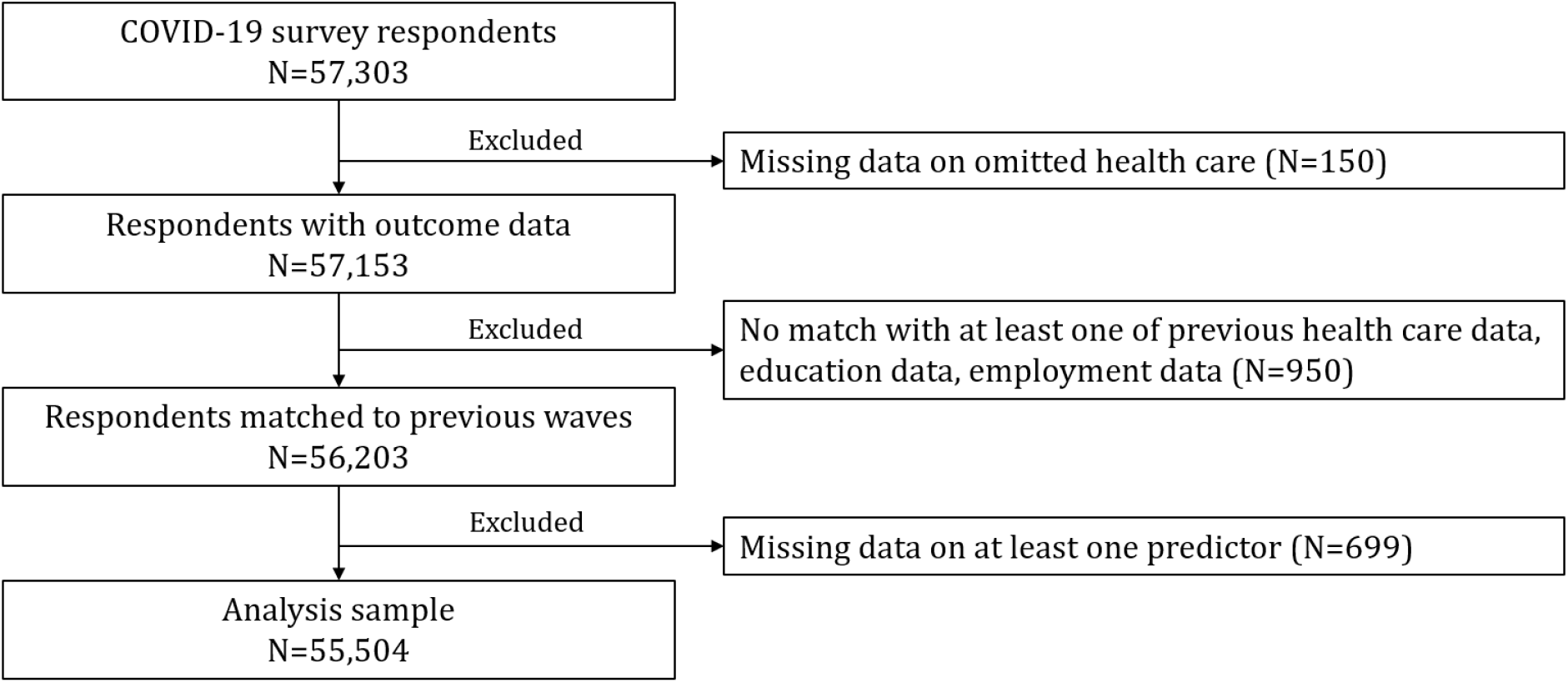
Sample selection process. Note: From the analysis sample, four individuals were dropped randomly to allow for five evenly sized folds for cross-validation.

### Statistical analyses

We compared four different algorithms to select models to predict missed health care visits: stepwise selection (R package step), group lasso (R package grpreg), random forest (R package ranger), and neural networks (R package keras). This conceptually involved two steps. First, we identified which of the many available predictors should be included in the predictive models (known as feature selection). Second, we compared different ways of using these predictors – through different model types and algorithms – to predict missed visits.

Stepwise selection and group lasso approaches combine both feature selection and prediction. Since our outcome is a binary indicator of whether an individual missed a visit, both approaches used logistic regression models for prediction. Therefore, any difference in prediction between the stepwise and group lasso approaches is purely due to differences in which predictors were selected for inclusion. For stepwise selection, the algorithm fits models of different sizes, sequentially deciding which predictor to add or drop to improve model performance. Among the many candidate models, the model with the highest Akaike Information Criteria is chosen. For group lasso, a penalty on the number of predictors is added to the logistic regression optimization, such that predictors with minimal predictive power are excluded.^25,26^ The optimal size of the penalty is defined as the penalty which minimizes the predictive error rate.

Both stepwise and group lasso approaches are appealing because they are based around logistic regression models – and thus are straightforward to interpret – and because they provide a principled way of deciding which predictors to include. However, they are limited in that they require the analyst to specify the functional form of each predictor (e.g. should age be included as a linear, quadratic, or cubic function?) and which interactions between predictors should be included. Random forests and neural networks, in contrast, do not explicitly decide which predictors to include, but they do identify which interactions and functional forms maximize predictive power. Random forests are based on decision trees.^27^ Decision trees are built by splitting the sample predictor by predictor such that the resulting sub-samples are more homogeneous regarding the outcome of interest. This results in a “tree”, where every split point (referred to as node) is essentially a decision rule on how to classify individuals based on the value of a predictor. We use the proportion of “positive” outcomes (i.e., missed health care visits) in a terminal subsample to estimate the probability of missed health care visits for a given individual (“probability tree”).^28^ Neural networks flexibly link predictors over several steps, known as “hidden” layers. In the simplest form (i.e., one “hidden” layer), linear functions with different weights for the predictors are combined to predict the outcome of interest. Then, the algorithm compares the predicted with the actual outcomes and adjusts the weights of the functions and the weights of the predictors within the functions to optimize the prediction. In more complex neural networks, more layers are added which combine the functions of the previous layer into new functions, thus allowing for more flexible interactions of the predictors.

The predictive power of group lasso, random forests and neural networks depend on the choice of specific parameters. To avoid overfitting the data and to ensure out-of-sample predictive validity, we choose the parameters using cross-validation. This involves splitting the data into several random subsets (known as folds), and using all folds but one to estimate the model. The model is then applied to the remaining fold to measure its out-of-sample performance. This procedure is repeated until every fold has been held out as a validation set. The specific characteristics we estimate through cross-validation are the penalty parameter for the group lasso, the optimal number of trees and the number of predictors among which the algorithm can choose to split the sample at each node for random forests, and the layer sizes and weights for neural networks. The final model parameters are depicted in the supplementary material.

Relatedly, the characteristics of each model depend on the input data, such that we assessed the performance of the final models using 5-fold cross-validation as well. Within each fold, we note the predictive accuracy (the proportion of correctly classified individuals), the sensitivity or true-positive rate (the proportion of individuals who were correctly classified among those who missed a care visit), and the specificity or true-negative rate (the proportion of individuals who were correctly classified among those who did not miss a care visit). We then graph these rates and average them over the five cross-validation folds for each algorithm.

All the models generate a predicted probability of missing a visit for each individual in the data; whether the individual is classified as having missed a visit depends on whether an individual’s predicted probability falls above or below a pre-specified probability cutoff. For all our results, we present the predictive performance measures across the entire range of potential probability cutoffs. This allows potential users to decide which cutoff meets their needs and resource constraints (e.g. a lower cutoff will likely have a higher true-negative rate but at the cost of resources needed to contact more individuals).

Next, we assess the importance of each predictor. For stepwise selection and group lasso, the measure of importance was the coefficient estimates from a logistic regression run on the full sample, including the predictors selected in the majority of the cross-validation folds. For random forest, we measured the importance of each predictor as the mean decrease in the Gini impurity.

All analyses were conducted in R 4.1.2.

## Results

### Sample characteristics

### Error! Reference source not found

displays the unweighted sample characteristics. There are more women than men in the sample (58% vs. 42%). Only a few respondents are younger than age 50 (<1%); these respondents are partners of the main SHARE respondents. More than half of the respondents (56%) are between 60 and 74 years old. Correspondingly, most are retired (64%). More than two-thirds completed at least upper secondary education (71%). Based on the previous diagnoses and medication, the majority of respondents require some form of regular care, especially for cardiovascular diseases such as hypertension (diagnosed: 56%, past medication: 56%) or high cholesterol (diagnosed: 41%, past medication: 37%). Every sixth respondent reported some type of missed essential health care (16%). More specifically, 7% reported they forwent care due to fear of COVID-19, 10% that the medical staff postponed treatment due to COVID-19, and 2% that they were denied health care (multiple answers possible).

### Prediction accuracy

Figure 2 displays the predictive accuracy of the four machine learning approaches. Each graph shows three different measures: (1) the true positive rate (sensitivity) (2) true negative rate (specificity) and (3) accuracy. The x-axis for each graph shows the cutoff point that is used to decide whether an individual’s predicted probability of a missed visit is classified as missed visit.

**Figure 2.**
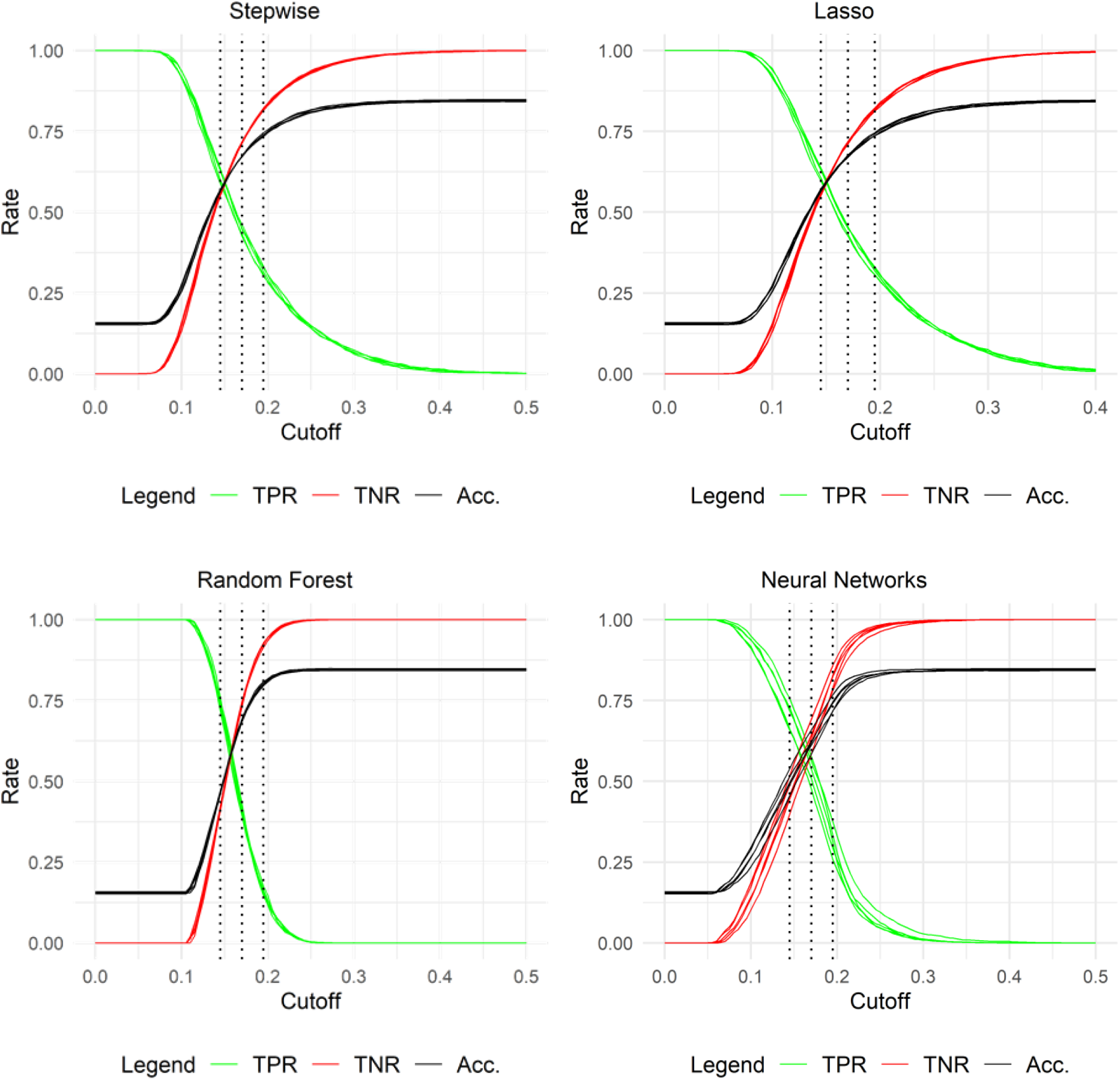
Performance measures Note: “TPR” is the true positive rate, “TNR” the true negative rate, and “Acc.” the accuracy. The outcomes for all five cross-validation steps are displayed. The cutoffs on the x-axis denote the predicted probability from which onwards individuals are labelled as “positive”, i.e. at risk of missing care.

Stepwise selection and group lasso perform very similarly. Starting at a cutoff of approximately 0.08, the true positive rate starts to decrease and the true negative rate and the accuracy start to increase. This indicates that higher cutoffs increase accuracy by classifying a greater percentage of individuals as not at risk with the trade-off that fewer individuals that missed visits are correctly classified as at risk. This trend stops at cutoffs above 0.4. The neural network models have a similar relationship between the probability cutoff and accuracy measures. In contrast, the random forest algorithm produces much steeper curves, with a sharp decline in the true positive rate and a sharp increase in the true negative rate and accuracy between cutoffs of 0.1 and 0.25. This indicates that the predicted probabilities of a missed health care visit mostly lie within this range and are thus more densely distributed compared to the other methods (eFigure 1 in the supplementary material displays the predicted probabilities for each method and confirms this interpretation). Overall, this implies that for random forest, small changes in the cutoff come with relatively larger changes in the share of targeted individuals. The other three methods are less sensitive to changes in the cutoff.

Table 2 compares the performance of the four methods at three different cutoffs to take a closer look at how the cutoffs can be used to decide on a targeting strategy. The optimal cutoff for targeting interventions depends on the trade-off the practitioner is willing to take between reaching out to the individuals at risk and falsely targeting individuals not at risk. As the distribution of predicted probabilities differs across the methods, the optimal cutoff also depends on the choice of the method. When classifying all individuals with a predicted probability of missed care visits above 14.5% as at risk, the algorithms can classify between 62% (stepwise selection and group lasso) and 74% (random forest) correctly, albeit at the costs of misclassifying between 44% (stepwise selection and group lasso) and 58% (random forest) of the individuals not at risk, with neural networks between them (70% vs. 54%). As the majority of individuals in the sample are not at risk, this implies that a large share of the thus targeted individuals would not be actually at risk. Thus, while correctly identifying many of the individuals at risk, such a strategy would be quite cost-intensive per correctly targeted individual. In contrast, targeting all individuals with a predicted probability above 19.5% entails targeting only between 16% (random forest) and 32% (stepwise selection, group lasso) of the individuals at risk, but would in turn only include between 8% (random forest) and 18% (stepwise selection, group lasso) of the population not at risk. In-between, at a cutoff of 17%, between 41% (random forest) and 53% (neural networks) of the individuals at risk could be reached, while falsely targeting between 36% (neural networks) and 26% (random forest) of the individuals not at risk.

To compare the performance of all four methods independent of the choice of cutoff, we display the area under the curve (AUC) in the last row of Table 2. This is the area under the curve when plotting the true positive rate on the false positive rate (1 -true negative rate). A method that would perform no better than chance would have an AUC of 0.5, an ideal method would have an AUC close to 1. Using this metric, all four methods perform very similarly with AUCs around 0.62. The results are qualitatively the same when disaggregating missed essential care visits into forgone visits due to fear, postponements by the medical staff, and denied care by the medical staff (eTable 1).

**Table 1.**
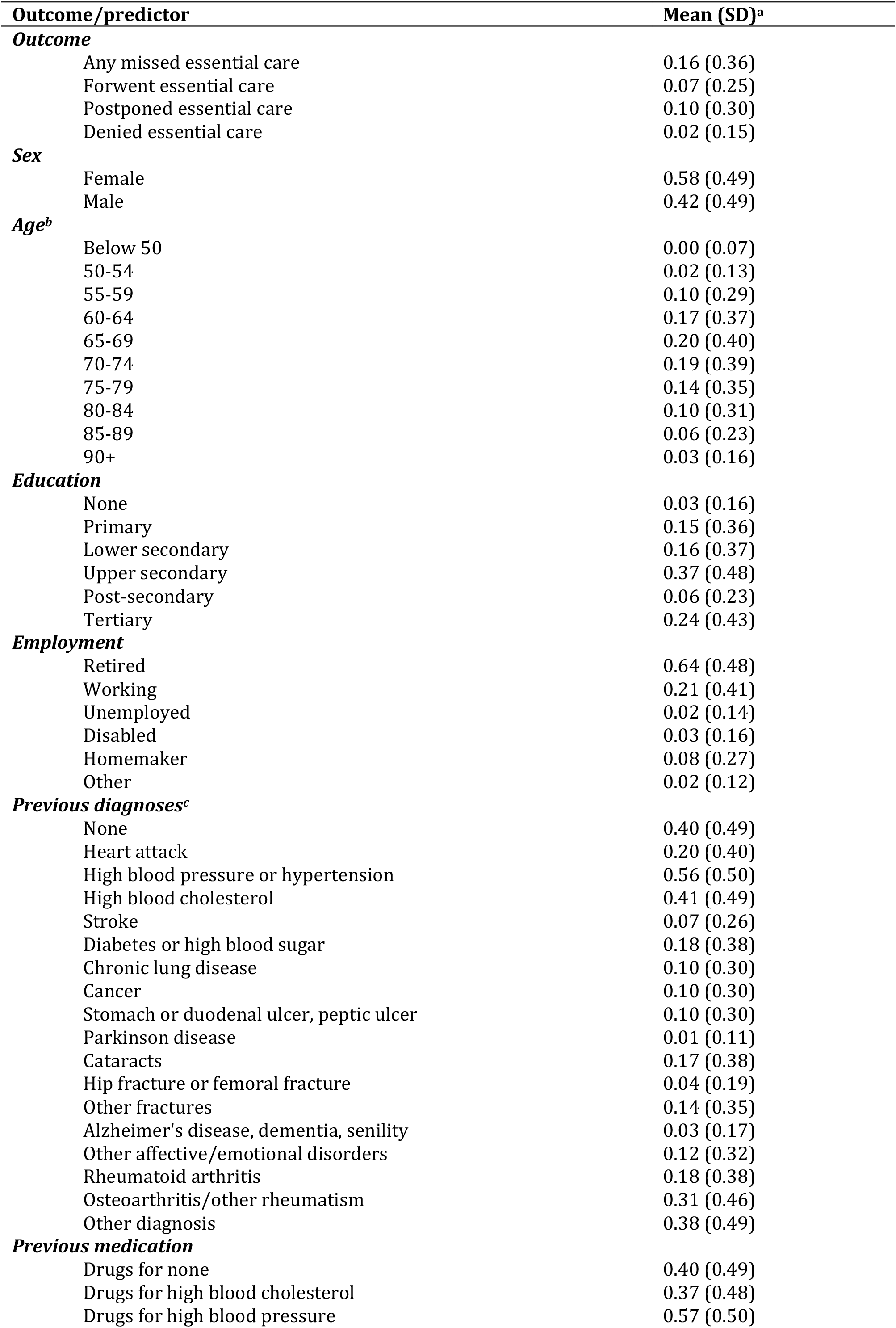
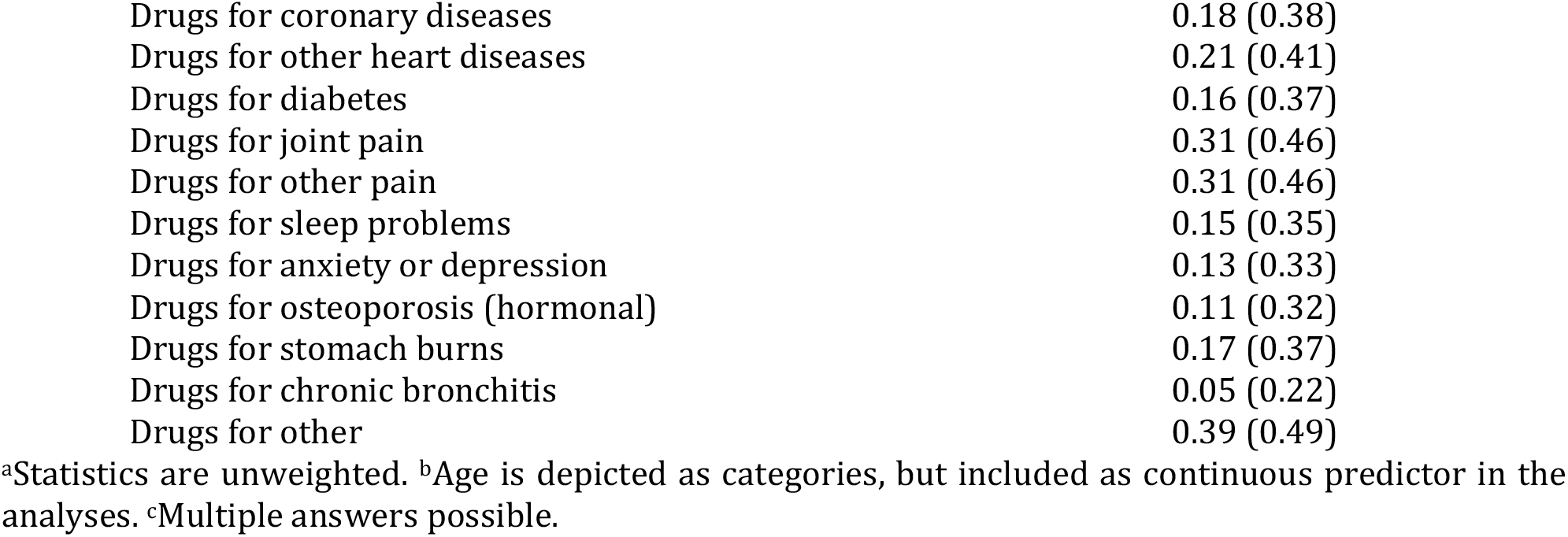
Sample characteristics.

| **Outcome/predictor** |  | **Mean (SD)<sup>a</sup>** |
| --- | --- | --- |
| ***Outcome*** |  |  |
|  | Any missed essential care | 0.16 (0.36) |
|  | Forwent essential care | 0.07 (0.25) |
|  | Postponed essential care | 0.10 (0.30) |
|  | Denied essential care | 0.02 (0.15) |
| ***Sex*** |  |  |
|  | Female | 0.58 (0.49) |
|  | Male | 0.42 (0.49) |
| ***Age<sup>b</sup>*** |  |  |
|  | Below 50 | 0.00 (0.07) |
|  | 50-54 | 0.02 (0.13) |
|  | 55-59 | 0.10 (0.29) |
|  | 60-64 | 0.17 (0.37) |
|  | 65-69 | 0.20 (0.40) |
|  | 70-74 | 0.19 (0.39) |
|  | 75-79 | 0.14 (0.35) |
|  | 80-84 | 0.10 (0.31) |
|  | 85-89 | 0.06 (0.23) |
|  | 90+ | 0.03 (0.16) |
| ***Education*** |  |  |
|  | None | 0.03 (0.16) |
|  | Primary | 0.15 (0.36) |
|  | Lower secondary | 0.16 (0.37) |
|  | Upper secondary | 0.37 (0.48) |
|  | Post-secondary | 0.06 (0.23) |
|  | Tertiary | 0.24 (0.43) |
| ***Employment*** |  |  |
|  | Retired | 0.64 (0.48) |
|  | Working | 0.21 (0.41) |
|  | Unemployed | 0.02 (0.14) |
|  | Disabled | 0.03 (0.16) |
|  | Homemaker | 0.08 (0.27) |
|  | Other | 0.02 (0.12) |
| ***Previous diagnoses<sup>c</sup>*** |  |  |
|  | None | 0.40 (0.49) |
|  | Heart attack | 0.20 (0.40) |
|  | High blood pressure or hypertension | 0.56 (0.50) |
|  | High blood cholesterol | 0.41 (0.49) |
|  | Stroke | 0.07 (0.26) |
|  | Diabetes or high blood sugar | 0.18 (0.38) |
|  | Chronic lung disease | 0.10 (0.30) |
|  | Cancer | 0.10 (0.30) |
|  | Stomach or duodenal ulcer, peptic ulcer | 0.10 (0.30) |
|  | Parkinson disease | 0.01 (0.11) |
|  | Cataracts | 0.17 (0.38) |
|  | Hip fracture or femoral fracture | 0.04 (0.19) |
|  | Other fractures | 0.14 (0.35) |
|  | Alzheimer's disease, dementia, senility | 0.03 (0.17) |
|  | Other affective/emotional disorders | 0.12 (0.32) |
|  | Rheumatoid arthritis | 0.18 (0.38) |
|  | Osteoarthritis/other rheumatism | 0.31 (0.46) |
|  | Other diagnosis | 0.38 (0.49) |
| ***Previous medication*** |  |  |
|  | Drugs for none | 0.40 (0.49) |
|  | Drugs for high blood cholesterol | 0.37 (0.48) |
|  | Drugs for high blood pressure | 0.57 (0.50) |

|  |  |
| --- | --- |
| Drugs for coronary diseases | 0.18 (0.38) |
| Drugs for other heart diseases | 0.21 (0.41) |
| Drugs for diabetes | 0.16 (0.37) |
| Drugs for joint pain | 0.31 (0.46) |
| Drugs for other pain | 0.31 (0.46) |
| Drugs for sleep problems | 0.15 (0.35) |
| Drugs for anxiety or depression | 0.13 (0.33) |
| Drugs for osteoporosis (hormonal) | 0.11 (0.32) |
| Drugs for stomach burns | 0.17 (0.37) |
| Drugs for chronic bronchitis | 0.05 (0.22) |
| Drugs for other | 0.39 (0.49) |

<sup>a</sup>Statistics are unweighted. <sup>b</sup>Age is depicted as categories, but included as continuous predictor in the analyses. <sup>c</sup>Multiple answers possible.

**Table 2.** Performance at three selected cutoffs.

| Performance measures | Stepwise | Lasso | Random Forest | Neural Networks |
| --- | --- | --- | --- | --- |
| ***Cutoff 1: 0.145*** |  |  |  |  |
| Accuracy | 0.5685 | 0.5692 | 0.4672 | 0.4993 |
| True positive rate | 0.6166 | 0.6203 | 0.7403 | 0.7045 |
| True negative rate | 0.5597 | 0.5598 | 0.4168 | 0.4612 |
| ***Cutoff 2: 0.170*** |  |  |  |  |
| Accuracy | 0.6728 | 0.6740 | 0.6886 | 0.6195 |
| True positive rate | 0.4428 | 0.4437 | 0.4059 | 0.5319 |
| True negative rate | 0.7152 | 0.7165 | 0.7408 | 0.6354 |
| ***Cutoff 3: 0.195*** |  |  |  |  |
| Accuracy | 0.7398 | 0.7401 | 0.8012 | 0.7381 |
| True positive rate | 0.3177 | 0.3174 | 0.1592 | 0.3136 |
| True negative rate | 0.8176 | 0.8180 | 0.9196 | 0.8163 |
| ***AUC*** | 0.6205 | 0.6207 | 0.6180 | 0.6202 |

All measures are means across the five cross-validation folds for each method.

### Importance of the predictors

Figure 3 depicts the coefficient estimates for the stepwise selection and group lasso model (Panel A) as well as the importance measures for random forest (Panel B). Although stepwise selection and group lasso are feature selection algorithms, for these data, the algorithms included nearly all the predictors we considered. The differences in the coefficient estimates between these two methods is due to the small differences in which exact predictors were included. One notable difference is the inclusion of the female indicator in the group lasso, but not the stepwise selection model, which instead includes a sex-specific linear age trend, with both predictors being significantly different from null in the respective model. Also, the group lasso model includes sex-specific differences in employment status, and more health-related predictors than the stepwise selection model. Yet, these additionally included predictors are not significantly different from zero on any conventional level. This is in line with the finding that both methods perform very similar in the prediction. Many predictors seem to be equally important, but two major trends stand out: Firstly, there seem to be substantial gender differences, either expressed as level effects (as in the group lasso), or through different age trends between men and women (as in both models). Secondly, a worse health condition, proxied by a disability to work or the medical history, increases the likelihood to report a missed health care visit. The only strong deviation is the presence of Alzheimer’s disease or a similar condition, which decreases the likelihood. This might be due to a higher probability to live in a nursing home and thus a higher care intensity, or because the answers of these respondents are more likely to be given either partially or completely by a proxy (40% of cases, versus 6% for all other respondents).

**Figure 3.**
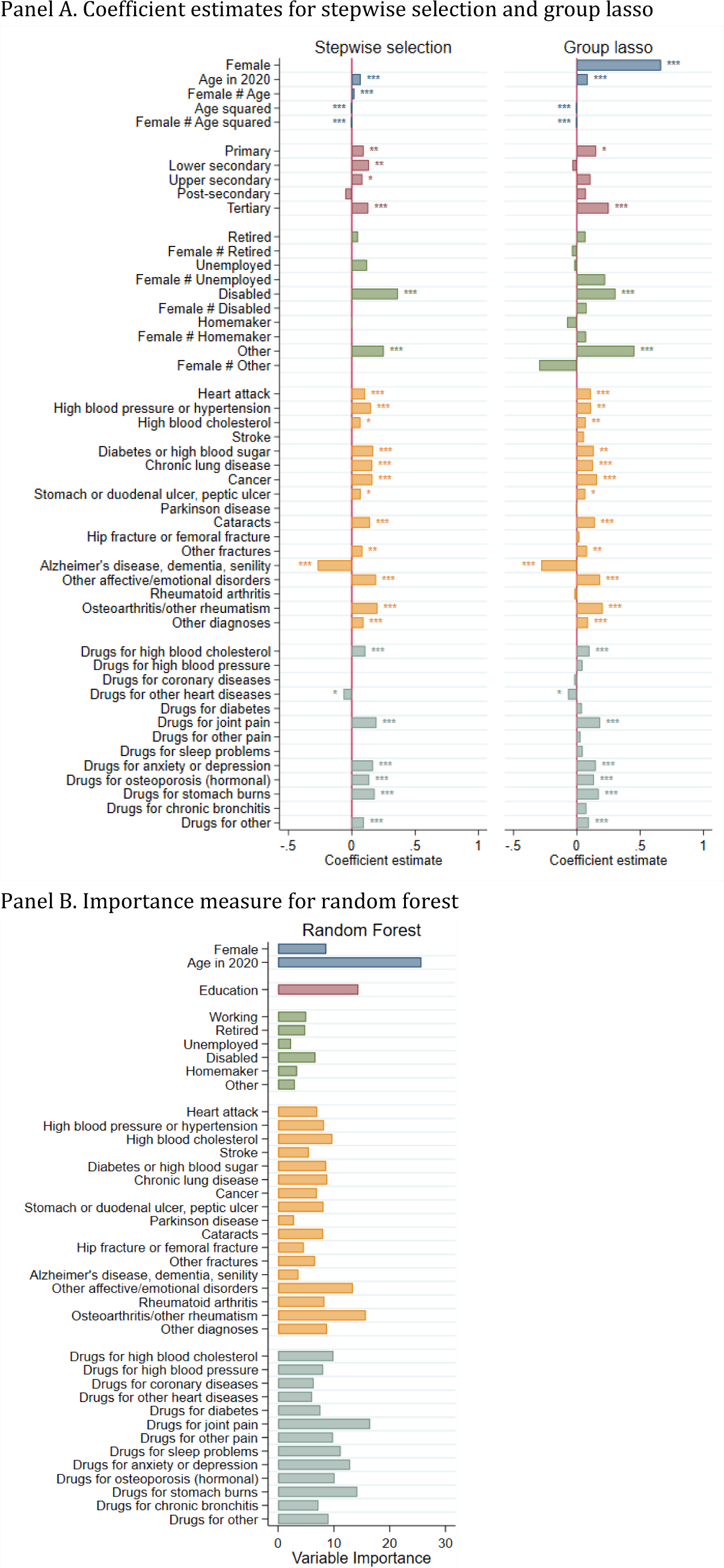
Coefficient estimates and importance measure. Note: Coefficient estimate for stepwise selection and group lasso stem from logistic regressions for the complete sample using only predictors selected by these algorithms. For random forest, the importance of predictors is the mean decrease in Gini impurity. Significance star defined as * p<0.1, ** p<0.5, ***p<0.01.

For random forest, the importance of the predictors is expressed as decrease in node impurity, and is positive by definition. Predictors with a higher value have a larger contribution to the decrease in node impurity over all trees. This measure tends to favor continuous predictors and categorical predictors with many categories, which is reflected in the high importance measure for age and education. Apart from these two predictors, a previous diagnosis of emotional disorders or osteoarthritis and a previous prescription of drugs for joint pain, stomach burns, or anxiety or depression have a high overall contribution.

## Discussion

In the light of repeated disruptions in health care over the ongoing waves of COVID-19, targeting individuals with missed essential health care visits is crucial to reduce the indirect impacts of COVID-19 on morbidity and mortality. Health insurance providers and similar stakeholders might be the natural choice for this, but might lack information on missed health care visits. To bridge this gap, repeatedly collected, large-scale surveys might be used to develop powerful predictive models which can be applied to health records data.

We used four different machine learning algorithms to predict missed health care visits: stepwise selection, group lasso, random forest and neural networks. All four methods perform quite similar. For example, when classifying all respondents with a predicted probability above 0.17 as at risk of a missed visit, both stepwise selection and group lasso had a true positive rate of 0.44, and a true negative rate of 0.72. This means that the predictions are correctly identifying 44% of those as at risk of a missed visit, and 72% of those not at risk. In comparison, the random forest had a slightly lower true positive rate (0.41), but a higher true negative rate (0.74), while the reverse was true for neural networks (0.53 and 0.64). The four methods differ in the spread of predicted probabilities: Random forest assigned probabilities in a very small range, resulting in densely distributed probabilities and hence a large sensitivity to changes in the cutoff.

Previous studies demonstrated that machine learning algorithms can be powerful tools to predict health care visits or no-shows.^29–34^ Their applicability in practice depends on the available data and the outcome of interest: For example, while data from Electronic Health Records (EHR) might contain a large number of predictors, including medical findings, health insurance providers might have access to claims data only.^35^ Still, a recent study showed that administrative data alone can be a powerful predictor for health care visits, with little additional improvement when adding further anthropometric data or information on neighborhood socioeconomic status.^30^ Similarly, non-attendance of appointments can be predicted very well based on previous appointments,^32–34^ but this data might not be available to health insurance providers. Moreover, even if such data is available, decisions to not visit a facility will not be recorded in administrative or clinical data. Using survey data, we show that missed health care visits can be predicted with machine learning data, but with a lower predictive power than reached by studies on health care visits or missed appointments.

Our predictions can be used to improve the targeting of individuals with missed health care visits during the first COVID-19 wave in Europe. The required weights and model parameters will be made available online, such that health insurance providers or similar stakeholders can estimate the probabilities of missed health care visits for their clients. Depending on the intended intervention, stakeholders can choose the cutoff which yields their preferred trade-off between reaching out to clients at risk, and falsely targeting clients not at risk. Moreover, as our analysis does not assess the potential negative consequences of missed health care visits, stakeholders might choose to weight our predictions of missed health care visits with their own assessments of severity of missed health care visits for specific subgroups, such as clients with a chronic illness.

Using survey data to predict missed health care visits is both a strength of our study and a potential bottleneck for the improvement of the predictions. To combine the benefits of survey data and health insurance provider data, we must focus on the information available in both datasets. This restricts us to a comparatively small set of predictors. Although the SHARE data already includes a comprehensive set of diagnoses and medication, future research could investigate whether extending these categories further would improve the predictive power for health care behavior sufficiently to balance the additional effort this entails for the survey administration. Moreover, data from new survey waves can help to understand how the predictive power of our models change over time, and thus, how applicable they are to target individuals throughout the pandemic.

Our study faces several limitations. First of all, our data relies on self-reported information on missed essential health care visits. This might introduce a bias if discrepancies between self-reported and objective missed health care visits are not random. In addition, participants were asked whether they had missed any health care visits since the onset of the COVID-19 pandemic. This might imply different recall periods for participants, as the onset of the pandemic is not a clear date and varies across locations. Still, given that nearly all European countries imposed the first lockdown within a time span of two weeks,^36^ and that this lockdown was an unparalleled, significant event, the resulting bias might be low. Similarly, most of the interviews took place within two months. While this might increase the recall period for the later participants, the interviews were conducted between the first and the second COVID-19 wave, in a time of comparatively low infection and death rates, such that most of the missed health care visits are expected to have already taken place before the start of the survey. Finally, we do not include missed health care at specialists, as the data combines specialists and dentists in one item. Given the high share of missed regular dentist check-ups recorded in other studies,^37,38^ we expect that these contribute to the majority of missed visits in this item, and thus are confident that excluding it leads to a more accurate identification of missed essential health care services.

The COVID-19 pandemic put health systems worldwide under pressure and led to severe disruptions in health care. With the support of machine learning methods, routinely-collected survey data can be used to target individuals at risk of missed health care more efficiently.

## Ethics approval

The SHARE study was approved by the Ethics Committee at the University of Mannheim (waves 1‐4) and by the Ethics Council of the Max‐Planck‐Society (waves 5‐8). Additionally, country-specific ethics committees or institutional review boards approved implementations of SHARE in the participating countries. All study participants provided informed consent.

## Data Availability

All data used in this study can be retrieved from the SHARE data repository (http://www.share-project.org/data-access.html). Code for replication is available upon reasonable request to the authors.

## Acknowledgments

This paper uses data from SHARE Waves 1, 2, 3, 4, 5, 6, 7 and 8 (DOIs: 10.6103/SHARE.w1.710, 10.6103/SHARE.w2.710, 10.6103/SHARE.w3.710, 10.6103/SHARE.w4.710, 10.6103/SHARE.w5.710, 10.6103/SHARE.w6.710, 10.6103/SHARE.w7.711, 10.6103/SHARE.w8.100, 10.6103/SHARE.w8ca.100), see Börsch-Supan et al. (2013) and Scherpenzeel et al. (2020) for methodological details.^15,39^ The SHARE data collection has been funded by the European Commission, DG RTD through FP5 (QLK6-CT-2001-00360), FP6 (SHARE-I3: RII-CT-2006-062193, COMPARE: CIT5-CT-2005-028857, SHARELIFE: CIT4-CT-2006-028812), FP7 (SHARE-PREP: GA N°211909, SHARE-LEAP: GA N°227822, SHARE M4: GA N°261982, DASISH: GA N°283646) and Horizon 2020 (SHARE-DEV3: GA N°676536, SHARE-COHESION: GA N°870628, SERISS: GA N°654221, SSHOC: GA N°823782) and by DG Employment, Social Affairs &Inclusion through VS 2015/0195, VS 2016/0135, VS 2018/0285, VS 2019/0332, and VS 2020/0313. Additional funding from the German Ministry of Education and Research, the Max Planck Society for the Advancement of Science, the U.S. National Institute on Aging (U01_AG09740-13S2, P01_AG005842, P01_AG08291, P30_AG12815, R21_AG025169, Y1-AG-4553-01, IAG_BSR06-11, OGHA_04-064, HHSN271201300071C, RAG052527A) and from various national funding sources is gratefully acknowledged (see www.share-project.org).

