## Supplementary material for "Predicting missed health care visits during the COVID-19 pandemic using machine learning methods: Evidence from 55,500 individuals from 28 European Countries"

#### Details on the machine learning methods

##### Stepwise selection

We implemented stepwise selection by using the function "step" in the package "stats" on a logistic function (using "glm"). We included education as ordered factor, employment and female employment (interaction between the female indicator and employment) as factors without ordering. We ran the function in a 5-fold CV to assess the predictive power. We selected the predictors which were chosen in the majority of the CV folds to construct a new model and run this on the complete data to obtain the coefficient estimates.

##### Group lasso

We implemented group lasso by using the function "cv.grpreg" in the package "grpreg". We specified a logistic function. We included education, employment and female employment as indicators for each level except the base levels and marked the indicators of each predictor as belonging to the same group such that the algorithm can drop either all indicators of one group or none. The "cv.grpreg" function internally implements a 10-fold CV procedure to determine the optimal regularization parameter lambda. As for stepwise selection, we ran the function in a 5-fold CV to assess the predictive power. We selected the predictors which were chosen in the majority of the CV folds to construct a new model and run this on the complete data to obtain the coefficient estimates.

##### Random forest

We implemented random forest by using the function "ranger" in the same-named package. We specified probability trees following Malley et al. (2012), such that classification trees were created, but the predicted outcomes were based on the share of positive cases in a node instead of a majority rule. This gave us predicted probabilities instead of predicted classes. We included employment and female employment as indicators for each level and education as ordered factor. We determined the optimal number of trees and the optimal number of predictors to select from at each split by running a 5-fold CV on each possible combination of the two parameters (trees: 100 to 500 in steps of 100, predictors: 1 to 10 in steps of 1) and choosing the combination with the highest average AUC across the five folds. The results are depicted below. We used the optimal parameters to assess the predictive power across the five folds. We calculated the importance of each predictor by averaging the mean decrease in node impurity across the five folds.

| | Number of trees | Number of predictors |
|---|---|---|
| Any missed visit | 500 | 1 |
| Forwent care | 500 | 1 |
| Postponed care | 300 | 1 |
| Denied care | 200 | 1 |

##### Neural networks

We implemented neural networks by using the "keras" package. We included education and employment as indicators for each level except the base levels. Age was standardized such that in ranges between 0 and 1 to match the range of all other predictors. We specified a model with two hidden layers. We determined the size of the hidden layers by running 5-fold CV on all possible combinations of layer sizes between 20 and 40 in steps of 5 for the first layer, and 15 to 40 in steps of 5 for the second layer, and choosing the combination with the highest AUC across the five folds. The results and further specifications of the model are depicted below. We compiled the model using the Adam optimizer on the two metrics binary cross entropy and AUC, with 30 epochs, a batch size of 100, and an internal split for validation during the training of 0.2.

| | Input layer | Hidden layer 1 | Hidden layer 2 | Output layer |
|---|---|---|---|---|
| Size | 42 | Determined with CV: | Determined with CV: | 1 |
| - Any missed visit | | - 20 | - 20 | |
| - Forwent care | | - 20 | - 15 | |
| - Postponed care | | - 20 | - 15 | |
| - Denied care | | - 35 | - 35 | |
| Activation | | Leaky ReLU | Leaky ReLU | Sigmoid |
| Dropout | | 0.5 | 0.5 | |

#### Supplementary figures

**eFigure 1. Distribution of predicted probabilities of missed health care visits**

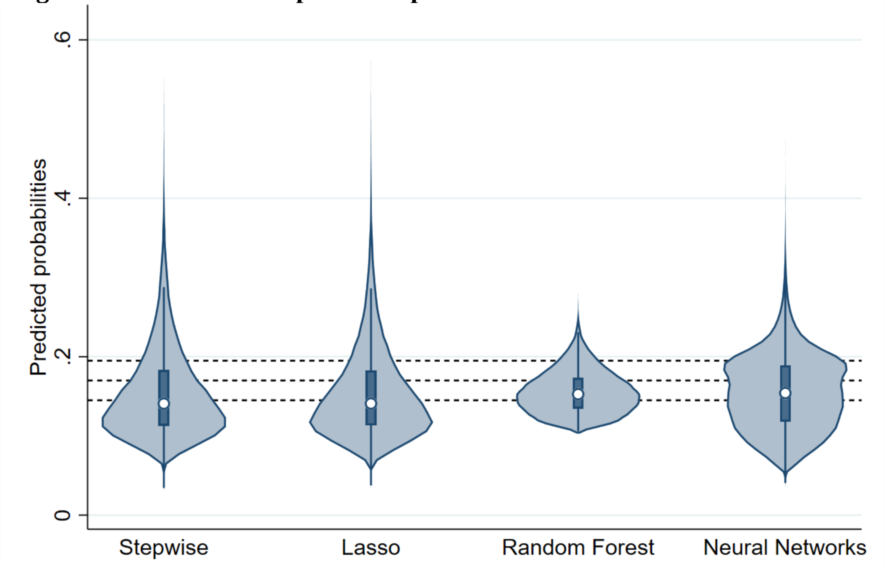


Note: Violin plots, with estimated kernel densities depicted as shaded area, the median as circle, the interquartile range as box and outliers as spikes. The dotted lines refer to the three cutoffs (0.145, 0.17, 0.195).

### Supplementary tables

**eTable 1. Model performance by disaggregated outcomes.**

| | | Stepwise select. | Group lasso | Random forest | Neural networks |
|---|---|---|---|---|---|
| **Forwent care due to fear** | **Cutoff 1** | 0.0600 | 0.0600 | 0.0600 | 0.0600 |
| | **Accuracy** | 0.5265 | 0.5282 | 0.4040 | 0.4292 |
| | **TPR** | 0.6466 | 0.6381 | 0.7754 | 0.7511 |
| | **TNR** | 0.5181 | 0.5205 | 0.3780 | 0.4067 |
| | **Cutoff 2** | 0.0700 | 0.0700 | 0.0700 | 0.0700 |
| | **Accuracy** | 0.6623 | 0.6656 | 0.6645 | 0.5660 |
| | **TPR** | 0.4899 | 0.4869 | 0.4795 | 0.6047 |
| | **TNR** | 0.6745 | 0.6782 | 0.6776 | 0.5633 |
| | **Cutoff 3** | 0.0800 | 0.0800 | 0.0800 | 0.0800 |
| | **Accuracy** | 0.7524 | 0.7570 | 0.8312 | 0.6867 |
| | **TPR** | 0.3653 | 0.3612 | 0.2211 | 0.4568 |
| | **TNR** | 0.7796 | 0.7848 | 0.8740 | 0.7028 |
| | **AUC** | 0.6139 | 0.6146 | 0.6160 | 0.6168 |
| **Medical staff postponed care** | **Cutoff 1** | 0.0800 | 0.0800 | 0.0800 | 0.0800 |
| | **Accuracy** | 0.4397 | 0.4340 | 0.2401 | 0.3801 |
| | **TPR** | 0.7516 | 0.7547 | 0.9228 | 0.8026 |
| | **TNR** | 0.4053 | 0.3986 | 0.1649 | 0.3334 |
| | **Cutoff 2** | 0.1000 | 0.1000 | 0.1000 | 0.1000 |
| | **Accuracy** | 0.6291 | 0.6293 | 0.5749 | 0.5581 |
| | **TPR** | 0.5262 | 0.5251 | 0.5871 | 0.6157 |
| | **TNR** | 0.6405 | 0.6408 | 0.5736 | 0.5516 |
| | **Cutoff 3** | 0.1200 | 0.1200 | 0.1200 | 0.1200 |
| | **Accuracy** | 0.7448 | 0.7469 | 0.8089 | 0.7146 |
| | **TPR** | 0.3529 | 0.3472 | 0.2283 | 0.3971 |
| | **TNR** | 0.7880 | 0.7909 | 0.8729 | 0.7495 |
| | **AUC** | 0.6178 | 0.6163 | 0.6143 | 0.6153 |
| **Medical staff denied care** | **Cutoff 1** | 0.0180 | 0.0180 | 0.0180 | 0.0180 |
| | **Accuracy** | 0.4699 | 0.4491 | 0.2399 | 0.5659 |
| | **TPR** | 0.6414 | 0.6751 | 0.8553 | 0.5854 |
| | **TNR** | 0.4661 | 0.4441 | 0.2263 | 0.5655 |
| | **Cutoff 2** | 0.0220 | 0.0220 | 0.0220 | 0.0220 |
| | **Accuracy** | 0.6702 | 0.6700 | 0.6363 | 0.7210 |
| | **TPR** | 0.4783 | 0.4876 | 0.5158 | 0.4071 |
| | **TNR** | 0.6744 | 0.6741 | 0.6390 | 0.7280 |
| | **Cutoff 3** | 0.0260 | 0.0260 | 0.0260 | 0.0260 |
| | **Accuracy** | 0.7794 | 0.7886 | 0.8323 | 0.8153 |
| | **TPR** | 0.3506 | 0.3402 | 0.2690 | 0.2990 |
| | **TNR** | 0.7889 | 0.7985 | 0.8448 | 0.8267 |
| | **AUC** | 0.5889 | 0.6003 | 0.6039 | 0.5991 |

All measures are means across the five cross-validation folds for each method.
